# The diagnostic value of plasma phosphorylated tau 181 in Alzheimer’s disease within the Chinese population: A systematic review and meta-analysis

**DOI:** 10.1101/2024.08.15.24312085

**Authors:** Keqiang Yan, Shuxin He, Xiaodong Jia, Haiyan Li, Dequan Liu, Jianchun Chen

**Affiliations:** Tianjin Kingmed Diagnostics Laboratory Co.Ltd, Tianjin, China; Tianjin Key Laboratory of Multi-omics Precision Diagnosis Technology for Neurological Diseases, Tianjin, China; Tianjin Medical University, Tianjin, China

**Keywords:** meta-analysis, p-tau181, Alzheimer’s disease

## Abstract

Among all related biomarkers, plasma phosphorylated tau (p-tau181) has demonstrated strong diagnostic performance in the very early stages of Alzheimer’s disease (AD), showing significant differences between AD patients and healthy controls. The aim of our present systematic review and meta-analysis was to roundly evaluate the clinical diagnostic value of plasma p-tau181 based on the Simoa platform in Chinese populations. We systematically searched five databases (Embase, PubMed, Cochrane Library, MEDLINE, and Web of Science) from inception to May 11th, 2024, as well as the references of retrieved relevant articles. We included prospective cohort studies and retrospective case-control studies in our analysis. Out of 1165 identified articles, 10 met the inclusion criteria for meta-analysis. Our quantitative analysis showed that plasma p-tau181 levels were significantly increased in patients with AD and mild cognitive impairment (MCI) compared to healthy controls (standard mean difference [SMD]: 1.45 [1.25 – 1.65], p<0.00001; SMD: 0.55 [0.31 – 0.78], p<0.00001) and were lower in MCI patients compared to AD patients (SMD: -0.88 [-0.93 – -0.82], p<0.00001). The reference values for plasma p-tau181 were 4.48 [95% confidence interval (CI): 4.01 – 5.00] for AD patients, 2.86 [95% CI: 2.45 – 3.34] for MCI patients, and 2.09 [95% CI: 1.90 – 2.30] for healthy controls. The meta-analysis confirmed that plasma p-tau181 significantly increases from healthy controls to mild cognitive impairment (MCI) and then to AD in the Chinese population. We also provide reliable reference values for plasma p-tau181, which contribute to the early diagnosis of AD in Chinese clinical settings.

## Introduction

Alzheimer’s disease (AD) is the most prevalent neurodegenerative disorder and a leading cause of dementia worldwide[1]. First identified by Dr. Alois Alzheimer in 1901, AD has since posed significant challenges to the medical community, with no definitive cure currently available[2]. The 2011 National Institute on Aging-Alzheimer’s Association (NIA-AA) Diagnostic Criteria conceptualize AD as a continuum encompassing three stages: an asymptomatic phase (preclinical AD), a predementia phase (mild cognitive impairment due to AD), and a dementia phase (dementia due to AD)[3–5]. Pathophysiological changes in AD can commence 15 to 20 years prior to the manifestation of clinical symptoms, underscoring the critical importance of early diagnosis and timely intervention to potentially slow disease progression[6].

Recent studies have focused on the early detection of AD through the use of fluid biomarkers[7]. Tau proteins, which are microtubule-binding proteins, play an essential role in maintaining neuronal stability and morphology under normal conditions. The regional progression of brain atrophy in AD is closely associated with the accumulation of Tau proteins[8]. Hyperphosphorylated Tau (p-tau), in its full-length or truncated forms, represents a key pathological hallmark of AD, exhibiting over 70 post-translational modifications, including more than 40 phosphorylation sites. Various forms of p-tau can be measured in cerebrospinal fluid (CSF) and plasma[9]. Among these, p-tau181 is significantly elevated in the CSF of AD patients and is considered a critical biomarker for the disease[10]. The ability of p-tau181 to differentiate AD from other neurodegenerative conditions, such as frontotemporal lobar degeneration (FTLD-TDP) and chronic traumatic encephalopathy (CTE), is crucial for the accurate diagnosis of AD[11]. Furthermore, p-tau181 levels correlate with AD severity and predict disease progression, providing valuable insights for evaluating therapeutic efficacy and guiding treatment strategies. Recent findings indicate that p-tau181 have excellent discriminative performance in plasma, paving the way for non-invasive monitoring of AD[12–15].

Advancements in technology, such as the Single-molecule Array (Simoa), have enabled the detection of plasma p-tau181 at low concentrations[16]. The US FDA has granted breakthrough medical device clearance to this technology, recognizing it as a valuable tool for diagnosing AD through the detection of p-tau181 biomarkers in blood[17–18]. Currently, this technique is also widely used in the clinical diagnosis of AD in China. Despite the proven diagnostic value of plasma p-tau181, a large amount of literature with inconsistent conclusions has confused our understanding of this field[19–24]. There is a notable lack of accurate clinical reference values that can distinguish AD from healthy elderly individuals. Previous studies, including one by Ding et al., have attempted to address this by comparing the range of p-tau181 between AD patients and healthy individuals through meta-analysis[25]. However, these studies predominantly collected data from European and American populations, which may not be applicable to other populations due to quantitative differences in p-tau181 levels among different races and ethnicities. Howell et al. found that older African Americans had lower CSF levels of p-tau181 compared to older Caucasians, even after adjusting for demographic variables, genetic factors, and cognitive function[26]. This suggests that there may be race-associated differences in CSF p-tau181 marker, which also exist in plasma.

To address this gap, the primary objective of the present study is to conduct an extensive and comprehensive meta-analysis to determine the distribution of plasma p-tau181 levels in both healthy individuals and patients with AD and other neurodegenerative diseases within the Chinese population. Our aim is to provide more precise and reliable cut-off thresholds for clinical use, thereby improving the accuracy of AD and other neurodegenerative disease diagnoses and facilitating early screening in Chinese clinical settings.

## Methods

### Search strategy and study selection

This meta-analysis was conducted in accordance with the Preferred Reporting Items for a Systematic Review and Meta-analysis (PRISMA) guidelines. The analysis questions were designed according to the PICOS framework:

- Participants: clinically diagnosed AD patients;
- Intervention: evaluation of plasma p-tau181levels;
- Comparison: patients clinically diagnosed with non-AD dementia and cognitively unimpaired participants;
- Outcome: normal range of plasma p-tau181 levels in healthy Chinese populations and the efficacy of plasma p-tau181 in differentiating AD from cognitively unimpaired participants or non-AD dementia patients, based on plasma p-tau181 level discrepancies;
- Study design: original, observational cross-sectional and prospective studies.

A literature search was conducted across five databases (Embase, PubMed, Cochrane Library, MEDLINE and Web of Science) on the date of May 11th, 2024, using the following search terms: plasma phosphorylated tau, Alzheimer disease, AD, Chinese and Simoa. Candidate studies were included or excluded based on the following criteria: (1) full text publications written in English; (2) plasma p-tau181 levels measured using Simoa; (3) Study population consisting of Chinese individuals; (4) AD diagnosed according to the 2011 and 2018 NIA-AA criteria[27] or the 1984 NINCDS-ADRDA criteria[28]. Studies were excluded if non-quantitative methods were employed to assess biomarker levels in the samples.

### Data extraction and quality assessment

Data of interest were independently extracted by two authors (Shuxin He and Keqiang Yan). Study quality was evaluated using the modified version of the Newcastle-Ottawa Scale (NOS)[29]. A maximum of eight points per study was given according to the quality of population selection, group comparability and exposure assessment. Disagreements regarding study selection or data extraction were resolved by additional reviewers (Xiaodong Jia or Haiyan Li).

### Statistical analysis

A random-effects model meta-analysis was employed to calculate the pooled effect size across multiple studies[31]. The normal range of plasma p-tau181 and standardized mean difference (SMD) between dementia interest groups were represented in forest plots with 95% confidence intervals (CI)[31]. Tau^2 was used to estimate of the between-study variance in a random-effects meta-analysis. Statistical heterogeneity across studies was assessed using chi-squared statistic (Chi^2) and the I^2 index. According to the Cochrane handbook, I^2 values of 25%, 50%, and 75% represented low, moderate, and substantial inter-study variability, respectively[32]. The Wan et al. method was employed to estimate the mean and standard deviation (SD) when studies provided median and interquartile range (IQR)[33]. All calculations were conducted using R statistical software (version 4.3.0).

## Results

### Study inclusions and quality assessment

The search syntax yielded 1165 publications, of which 275 were duplicates and 878 were excluded following abstract screening. Of the 22 entries that were eligible for full-text assessment, 12 studies were excluded for not complying with the eligibility criteria: Two studies were not based on a Chinese population; five did not provide specific diagnostic data for p-tau181; one did not use Simoa assays for measurement; and four had duplicated published data for the same baseline cohort. Consequently, 10 studies were included in this meta analysis. The detailed selection process is illustrated by the PRISMA flow diagram in Figure 1. The modified NOS criteria for quality assessment of the studies indicated that the majority of them were of high-quality (Table 1). One study scored low on the quality assessment since it only contained healthy cohort and lacked biomarker diagnosis for inclusion criteria. The characteristics of the 10 studies, including the number of subjects, average age, female percentage, and plasma p-tau181 level are presented in Table 2. From the included studies, 7 studies compared blood biomarkers between AD and healthy controls (783 AD patients and 508 controls), while four of them also focused on mild cognitive impairment (MCI) comparing 368 MCI patients and 321 controls. Six studies paid extra attention to the performance of plasma p-tau181 between 649 AD patients and 413 non-AD dementia patients (non-ADD), including patients with cognitive disorders other than AD, such as frontotemporal dementia, vascular dementia, Lewy body disease, and Parkinson’s disease dementia.

**Figure 1.**
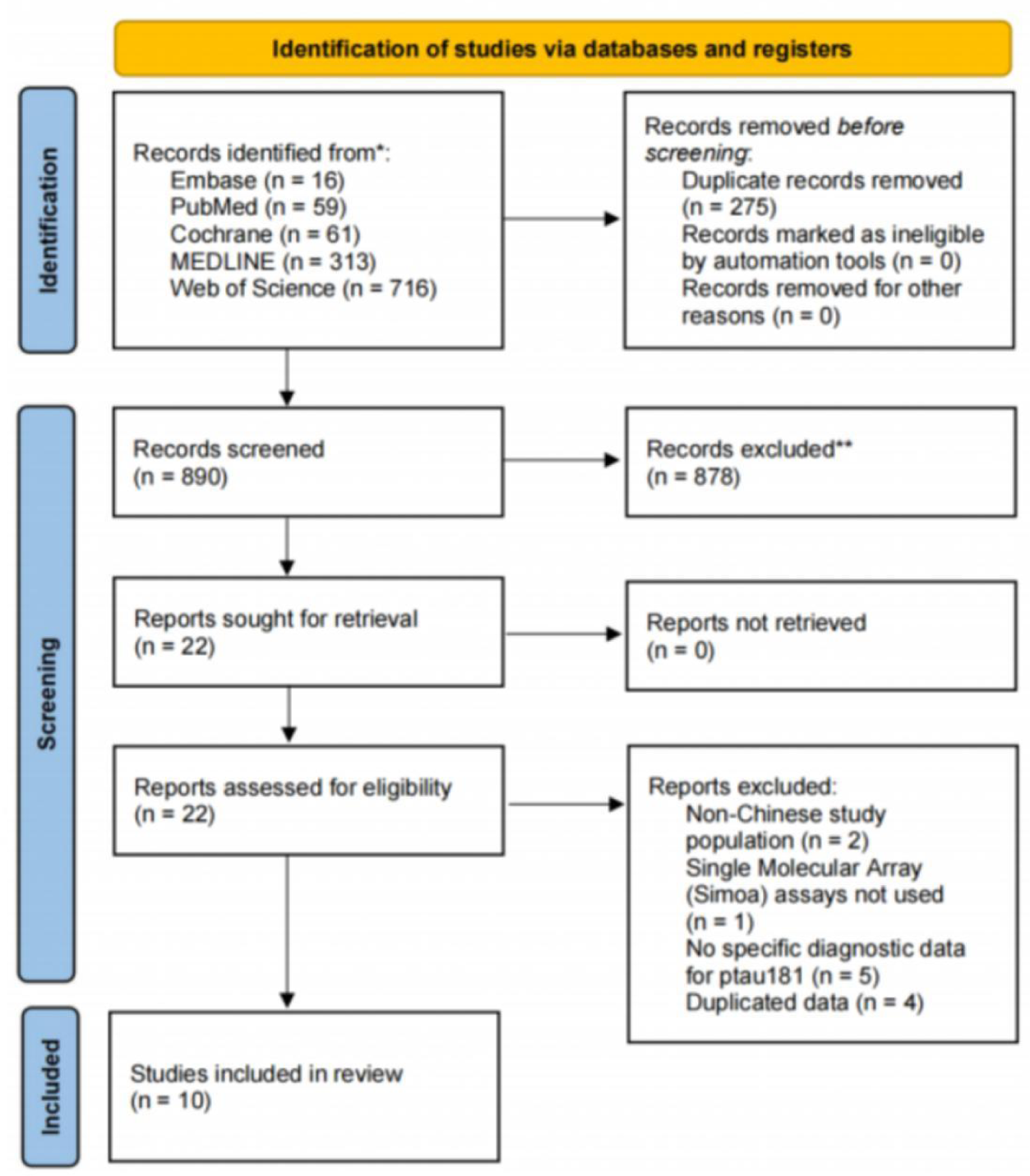
PRISMA flow diagram of study selection process

**Table 1.** Quality assessment of included studies in the meta-analysis using NEWCASTLE - OTTAWA QUALITY ASSESSMENT SCALE (NOS)

|  | Selection |  |  |  | Comparability | Exposure |  |  |
| --- | --- | --- | --- | --- | --- | --- | --- | --- |
|  | Is the case definition adequate? | Representativeness of the cases | Selection of Controls | Definition of Controls | Comparability of cases and controls on the basis of the design or analysis | Ascertainment of exposure | Same method of ascertainment for cases and controls | Non-Response rate |
| Chen J, et al. 2023 | NA | NA | - | - | NA | * | NA | * |
| Guo Y, et al. 2023 | * | * | * | * | ** | * | * | * |
| Sun Q, et al. 2022 | * | * | * | * | * | * | * | * |
| Cai H, et al. 2023 | * | * | * | * | * | * | * | * |
| Wu X, et al. 2021 | - | * | * | * | * | * | * | * |
| Gao F, et al. 2023 | * | * | * | * | * | * | * | * |
| Chen Y, et al. 2024 | * | * | * | * | * | * | * | * |
| Huang L, et al. 2024 | * | * | * | * | ** | * | * | * |
| Shang L, et al. 2023 | * | * | * | * | * | * | * | * |
| Xiao Z, et al. 2023 | - | * | * | * | * | * | * | * |

**Table 2.**
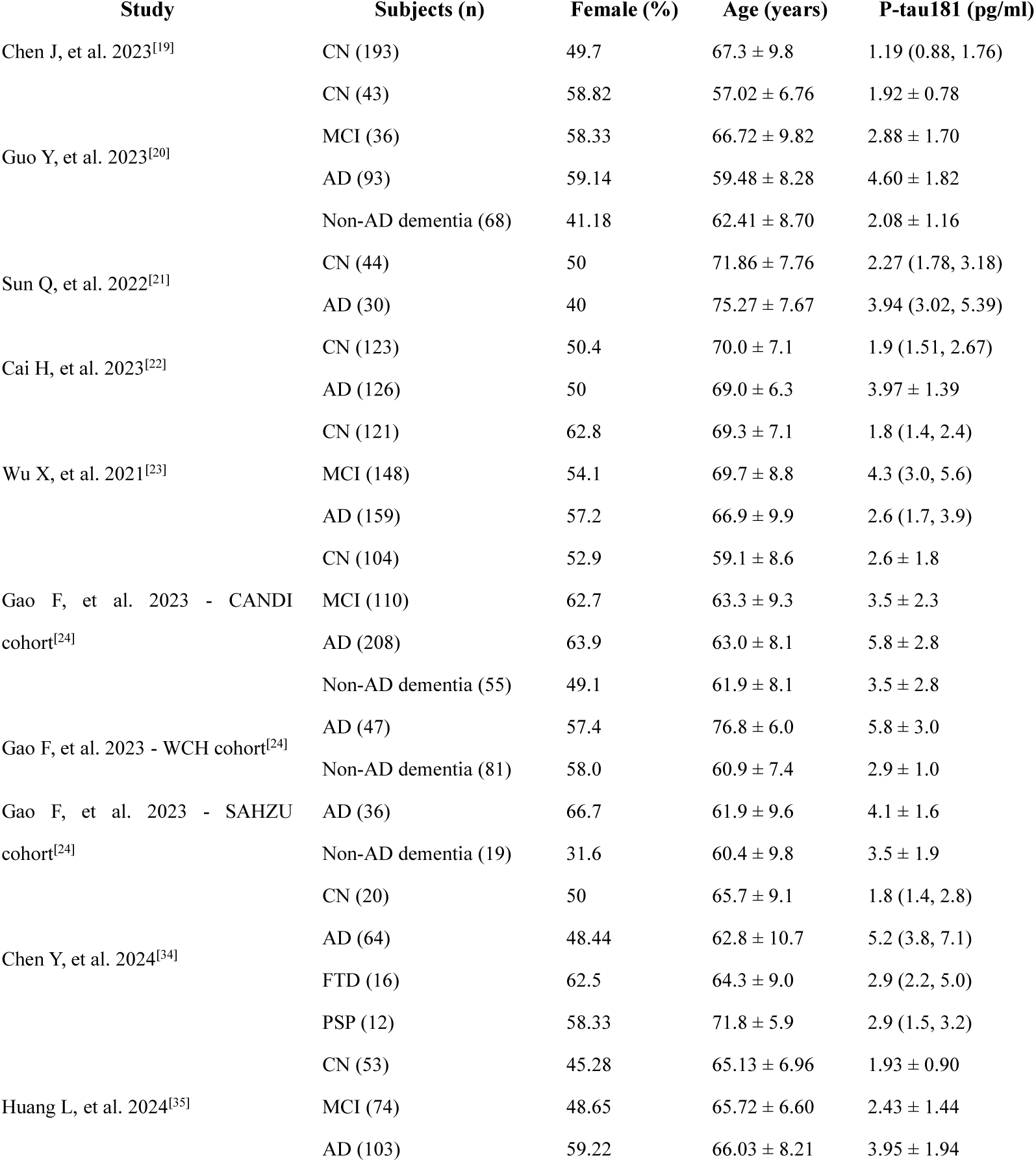
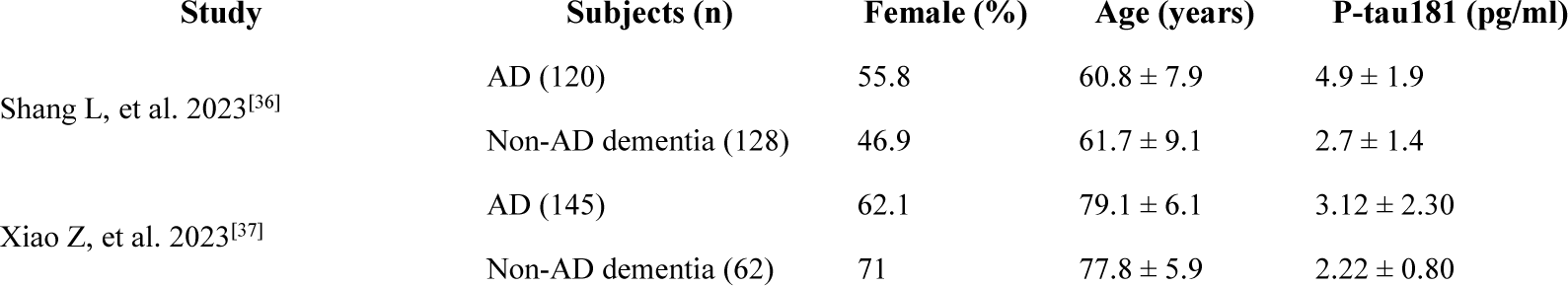
Basic characteristics of included studies for plasma p-tau181 analysis.

### Plasma p-tau181 levels in healthy cohorts

A summary of plasma p-tau181 levels in Chinese healthy cohorts was presented. Subgroup analysis was performed according to age and sex ratio. The retrieved data were stratified by mean age into two cohors: young (< 65 years) and old (≥ 65 years). The sex ratio was defined as medium (40-60% female) and high (more than 60% female). Following the removal of one study (Chen J, et al 2023) with high heterogeneity I^2 = 95%, seven studies were included, suggesting that the average plasma p-tau181 level in Chinese healthy populations was 2.09 pg/ml (95% CI 1.90–2.30, I^2 = 77%, P< 0.01) (Figure 2a). The mean plasma p-tau181 levels in young and old groups were 2.23 pg/ml (95% CI 1.66–3.00) and 2.03 pg/ml (95% CI 1.85–2.30), respectively, with no significant differences observed (p = 0.54) (Figure 2b). Nevertheless, there were significant differences (p = 0.04) in the subgroup analyses by sex ratio. As illustrated in Figure 2c, the mean plasma p-tau181 concerntration was 1.87 pg/ml (95% CI 1.74-2.01) in the subgroup with more than 60% female participants, in comparison with 2.14 pg/ml (95% CI 1.92-2.37) in the other subgroup.

**Figure 2.**
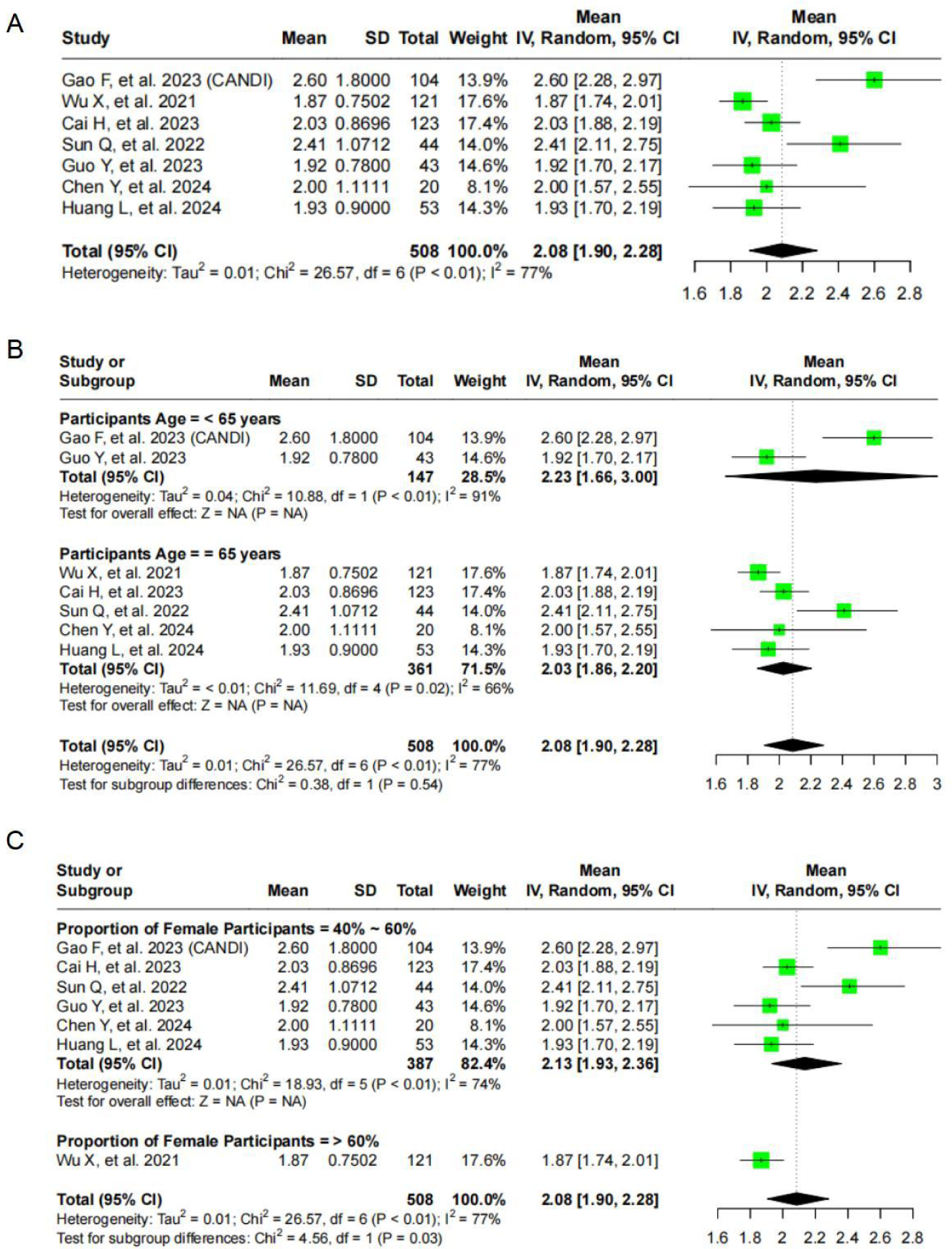
Forest plot of random effects meta-analysis of plasma p-tau181 levels in healthy controls (A), age subgroups (B) and sex ratio subgroups (C).

### Plasma p-tau181 levels in AD patients

A total of nine studies with 11 cohorts were conducted to evaluate p-tau181 plasma levels in 1131 AD patients. The results indicated a mean value of 4.48 pg/ml (95% CI 4.01-5.00, I^2 = 93%, P < 0.01) (Figure 3a). The heterogeneity of observed in the data was influenced by a number of factors, including age and sex composition, geographic location of the study, time period and the severity of disease. Most of the studies included in the current analysis employed clinical diagnostic guidelines, including various iterations of international guidelines, rather than the gold standard of autopsy-confirmed AD, which may have contributed to the observed heterogeneity. There was significantly difference of plasma p-tau181 levels between young and old groups of AD patients (P < 0.05), with 4.94 pg/ml (95% CI 4.42–5.53) and 4.12 pg/ml (95% CI 3.66–4.63) in young and old groups, respectively (Figure 3b). While no significant differences (P = 0.7) in subgroup analysis by sex ratio (Figure 3c). Subsequently, plasma p-tau181 levels were compared between AD patients and healthy cohorts. The meta-analysis based on seven studies including 783 AD patients and 508 control participants, demonstrated that p-tau181 was significantly higher in AD in comparison to control (p < 0.01), providing a SMD of 1.45 (95% CI: 1.25, 1.65), with low heterogeneity (I^2 = 33%) (Figure 3d). In the aspect of differentiating AD patients from normal controls, three studies demonstrated the diagnostic accuracy of plasma biomarkers, as indicated by ROC curves. Chen et al. reported as AUC (Area Under the Curve) of 0.882 (95%CI: 0.79, 0.97) for p-tau181, which was higher than Aβ40, Aβ42 and GAFP, but was lower than Aβ42/Aβ40 (AUC = 0.998, 95%CI: 0.99-1.00). Sun et al. combined plasma markers including Aβ42/Aβ40, p-tau181 and neurofilament light protein (NfL), suggesting the best discriminative performance (AUC = 0.902) among of all single plasma biomarkers (p-tau181 AUC=0.773), with a sensitivity and specificity of 0.867 and 0.886, respectively. Wu et al. found that p-tau181 performed well in both early-onset AD (EOAD) and late-onset AD (LOAD) patients (AUC = 0.925 and 0.842, respectively) for the AD/control comparison. Three additional studies focused on the ability of p-tau181 plasma levels in Aβ PET scanning status prediction, with AUC values of 0.813 (Xiao Z, et al., 2023), 0.916 (Guo Y, et al., 2023) and 0.839 (Gao F, et al., 2023 - CANDI cohort). Gao et al. constructed a logistic regression (LR) model, which combined APOE-ε4, plasma p-tau181, and serum glial fibrillary acidic protein (GFAP), resulting in an improvement in the AUC for distinguishing Aβ-PET status from 0.839 to 0.915. Subsequently, the efficacy of this LR model was demonstrated in both the CANDI and multi-center cohorts, showing superior performance compared to any of the blood biomarkers. Guo et al. found other plasma biomarkers including Aβ40, Aβ42, Aβ42/Aβ40 and Nfl, had relatively lower discriminative accuracy than p-tau181. Furthermore, the study reported that once the CSF Aβ42/Aβ40 ratio was positive, plasma p-tau181 began to increase significantly and continued to increase throughout the course of preclinical AD.

**Figure 3.**
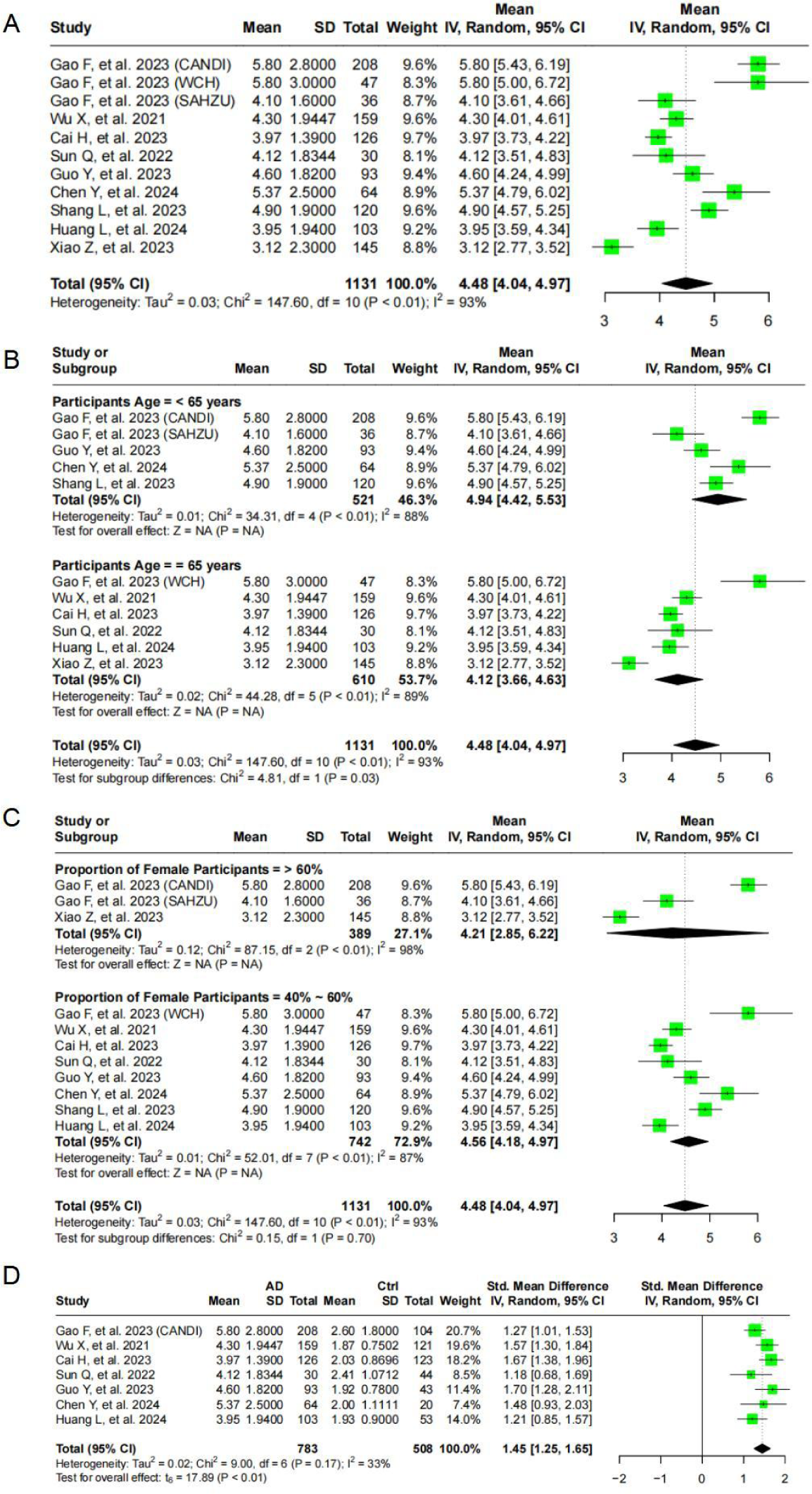
Forest plot of random effects meta-analysis of plasma p-tau181 levels in AD patients (A), age subgroups (B) and sex ratio subgroups (C). Meta-analysis of studies comparing p-tau181 levels between AD and healthy controls (D).

### Plasma p-tau181 in MCI patients

Four studies evaluated plasma p-tau181 levels in 368 patients with MCI, suggesting that the mean value was 2.86 pg/ml (95% CI 2.45–3.34, I^2 = 82%, P < 0.01) (Figure 4a). Our further meta-analysis revealed that p-tau181 levels were significantly higher in MCI patients compared with controls (SMD = 0.55, 95% CI: 0.31, 0.78, P < 0.01), while exhibiting a lower concentration than that in AD patients (SMD = -0.88, 95% CI: -0.93, -0.82, P < 0.01) (Figure 4b, 4c). One study (Wu X, et al. 2021) demonstrated that p-tau181 exhibited a larger AUC than plasma t-tau, Aβ40, Aβ42 and Nfl, achieving an AUC of 0.702 (95% CI: 0.64, 0.764) for the amnestic MCI/Ctrl comparison and 0.719 (95% CI: 0.661, 0.776) for the amnestic MCI/AD comparison. The AUC of the amnestic MCI/Ctrl comparison was then improved to 0.789 (95%CI: 0.737, 0.841) through the application of a model comparising t-tau, Aβ40, Aβ42, p-tau181, and NfL, in conjunction age, sex, and education years.

**Figure 4.**
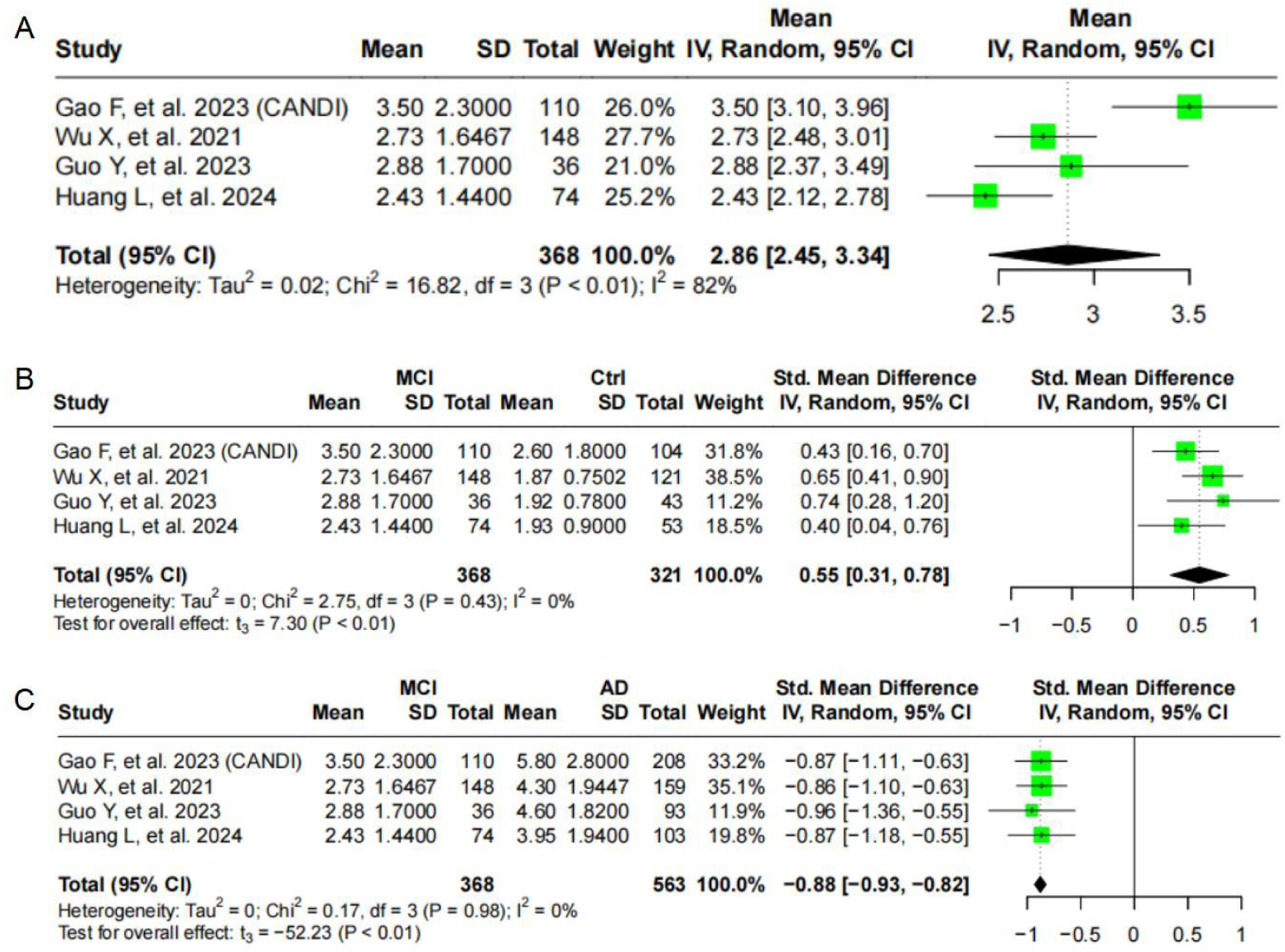
Forest plot of random effects meta-analysis of plasma p-tau181 levels in MCI patients (A), age subgroups (B) and sex ratio subgroups (C).

### Plasma p-tau181 between AD patients and non-AD dementia patients

Five studies with seven cohorts focused on the performance of plasma p-tau181 in distinguishing AD patients from non-AD dementia patients, demonstrating p-tau181 level was significantly higher in AD. Studies with small sample sizes were not included in the meta-analysis. Based on the meta-analysis results we proved that the average p-tau181 level in AD patients was significantly higher than in those non-ADD patients, providing a SMD of 1.12 (95% CI: 0.53,1.71, p<0.01) (Figure 5). Two studies reported that p-tau181 had a better performance in discriminating AD from non-ADD than Aβ42, Aβ42/40, GFAP and NfL, providing the AUC of 0.892 and 0.805 respectively[36–37]. Besides, the AUC was improved to 0.945 when combined with other plasma biomarker index (Aβ42/ Aβ40, NfL, GFAP) in Chen Y, et al. research.

**Figure 5.**
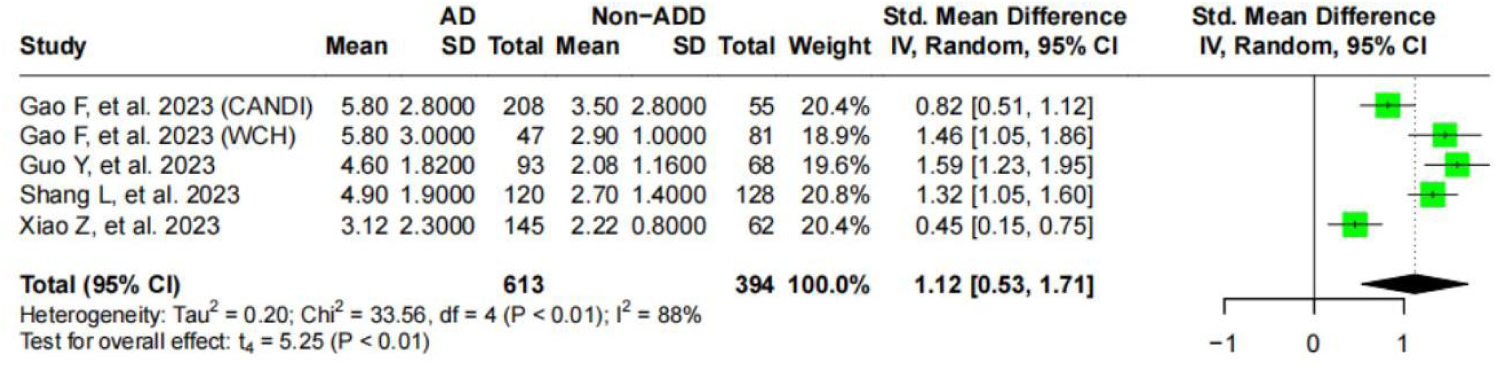
Forest plot of random effects meta-analysis of plasma p-tau181 levels in AD and non-AD patients.

## Discussion

The current investigation examined the conflicting results of studies on plasma p-tau181 linked to AD or MCI in the Chinese population, aiming to develop a non-invasive diagnostic biomarker for the early phases of AD. Plasma p-tau181 levels may serve as one of the most promising fluid biomarkers for validating AD or MCI diagnoses. The current findings show that AD is associated with higher blood levels of p-tau181 compared to MCI and controls. Based on the current literature, we established the average plasma p-tau181 levels for AD, MCI, and controls in the Chinese population, providing reference values for future clinical diagnosis.

Many meta-analysis studies have demonstrated that plasma p-tau181 can precisely differentiate AD from MCI and healthy controls. However, most of the studies were based on Western populations. It is validated that race and ethnicity have impact on the level of p-tau181 in many studies. Howell et al. found that African Americans showed lower p-tau181 levels in cerebrospinal fluid (CSF) than Caucasians, which may lead to underdiagnosis of AD in African Americans[26]. Schindler et al. Further validated that diagnosis performance of plasma p-tau181 in AD is inconsistent between African (AA) and non-Hispanic White (NHW), which could result in disproportionate misdiagnosis of AA individuals[38]. In the Canadian population, the reference range for plasma p-tau181 is 0.9-4 pg/ml for individuals aged 60-80. In a Japanese population study, the interquartile range (IQR) for plasma p-tau181 was 2.58-4.31 pg/ml in PET-positive cohorts, and 1.32-2.33 pg/ml in PET-negative individuals. Additionally, a meta-analysis by Ding et al. showed that the average plasma p-tau181 level in the general population is 11.18 pg/ml[25]. This is significantly different from the meta-analysis results for the Chinese population, which reported an average level of 2.09 pg/ml.This study was conducted among Chinese populations, which further add to the evidence of plasma p-tau181 as reliable prognostic candidates for AD and highlight their promising roles in future clinical practice and research. Cooper et al. found in addition to population, p-tau181 levels are also affected by other factors such as hypertension, myocardial infarction (MI) and stroke[39]. More comprehensive studies are needed to determine the impact of these factors on p-tau181 levels.

To improve diagnosis accuracy, the p-tau181 was combined with APOE genotyping, where the presence of an ε4 allele (encoding the E4 isoform of the apoplipoprotein E protein) is associated with higher probability of AD. Snellman et al. indicates that plasma p-tau181 levels are significantly higher in APOE4/4 carriers compared to non-carriers and heterozygous APOE3/4 carriers[40]. Elevated levels of plasma p-tau181 are already observed in cognitively unimpaired high-risk individuals with homozygous APOE4/4. Cooper et al suggests a strong correlation between p-tau181 levels and APOE ε4 carrier status in cognitively healthy individuals over 85 years old[39]. Although p-tau181 levels are elevated in elderly APOE ε4 carriers, their cognitive abilities remain intact. Combining APOE and p-tau181 markers and conducting joint analyses may provide more comprehensive predictive information for clinical diagnosis of Alzheimer’s disease.

Compared to p-tau181, p-tau217 which is tightly associated with Ab plaque formation within the brain, suggested a higher specificity to AD. Nervertheless, meta-analysis of this biomarker could not be done due to too few studies, especially in Chines population. Integration of diverse P-tau formed proteins such as p-tau181, p-tau217 and p-tau231 may significantly improved diagnosis power of AD[41].

### Limitations

Some limitations should be addressed in this meta-analysis. Firstly, misdiagnosis could have occurred in some of the studies included in this systematic review, as diagnosis was reached mostly on clinical grounds rather than gold-standard autopsy-confirmend AD. Secondly, only 10 studies were used for meta-analysis, this small studies can be more heterogeneous than large studies. These findings will be verified in future large and multicenter cohort studies for subsequent diagnosis for MCI and AD. Lastly, it is likely that the level of plasma p-tau181 is correlated with APOE status. However, due to lacking of these data, we can’t explore the impact of APOE to levels of p-tau181 in different groups.

## Conclusion

In summary, our meta-analysis revealed significant relationships of plasma p-tau181 with Alzheimer’s patients compared with cognitively normal control subjects and mild cognitive impairment. The results presented provide preliminary reference value of plasma p-tau181 for AD and MCI, which may facilitate clinical diagnosis of neurological diseases for Chinese population.

## Data Availability

All data produced are available online at

https://github.com/KeqiangYan/meta-analysis

## Authors’ contributions

KQ Yan conceptualised this systematic review. SX He searched the literature and screened the titles and abstracts. KQ Yan and SX He reviewed all full-text articles and extraced data. HY Li and XD Jia formally analysed, visualised and validated the data. DQ Liu and JC Chen supervised the systematic review. All authors were responsible for the methodology and review and editing of the manuscript. All authors had final responsibility for the decision to submit the manuscript for publication.

